# Co-occurrence Is Not Evidence: Separating Protein–Disease Findings from Incidental Mentions in HFpEF Literature

**DOI:** 10.64898/2026.11.21.26368635

**Authors:** Aktan Azat, Clodomir Joaquim De Santana Junior, Martin Cadeiras, David A. Liem

## Abstract

Literature-mining pipelines such as CaseOLAP rank proteins by how often they appear next to a disease term. Appearing next to a disease is not the same as being implicated in it: in our 300-sentence held-out set of PubMed sentences that mention a protein and heart failure with preserved ejection fraction (HFpEF), only 115 assert a biological association. The rest report a negative finding or mention the protein in passing, for example as a drug the patients were taking. We train a sentence-level filter that sorts each protein mention into associated, not_associated or incidental. It is a fine-tuned PubMedBERT trained with class-weighted cross-entropy and R-Drop regularization, combined into a six-model majority-vote ensemble over three random seeds. On the held-out set, with no thresholds tuned on it, the ensemble reaches 85.3% accuracy and 86.1% macro F1 (bootstrap 95% confidence intervals 81.0–89.3% and 82.1–89.7%), against 38.3% accuracy for the co-occurrence baseline, which counts every mention as an association. Single models average **84.4 ± 0.7%** accuracy across seeds. Run after CaseOLAP, the filter removes protein mentions that are not evidence of association from the HFpEF candidate list.

Where the ensemble’s majority vote disagreed with the annotation, the annotation was often wrong. Expert review of those sentences corrected 53 of 88 flagged evaluation labels and 109 training labels. Most of the errors came from one flaw in the annotation sheet: in sentences naming several proteins, it did not say which protein the label was for. The audit stage coincided with the largest accuracy gain in the project, but the same stage also changed the evaluation labels and the training loss, so we report it as a chronology, not as a controlled result.

## 1 Introduction

Heart failure with preserved ejection fraction (HFpEF) is heart failure in which the left ventricular ejection fraction stays at or above 50%. It accounts for roughly half of all heart failure cases worldwide and falls hardest on older adults and women (Redfield and Borlaug, 2023). Heart failure with reduced ejection fraction (HFrEF) is mainly a problem of pumping; HFpEF is mainly a problem of filling: the ventricle stiffens, diastolic filling suffers, and patients usually carry several comorbidities at once, most often obesity, hypertension, atrial fibrillation and chronic kidney disease.

HFpEF also has far fewer treatments. Sodium-glucose co-transporter 2 (SGLT2) inhibitors were the first drug class to show consistent benefit across large HFpEF trials, and the pathways behind that benefit are still partly unresolved. What is known about the proteins, biomarkers and signaling pathways that set HFpEF apart from HFrEF is scattered across thousands of PubMed abstracts from the past two decades, with no single place that collects it. Reviewing that literature by hand is slow, and relevant work gets missed.

Text mining is one way to collect it. Prior work has built knowledge graphs from co-occurrence statistics (Liem et al, 2018; Fang et al, 2021), applied biomedical language models to entity recognition and relation extraction (Lee et al, 2020; Gu et al, 2022), and built disease-specific filtering pipelines for cardiovascular targets (Bhatt et al, 2024; Urdang et al, 2025). These methods produce candidate protein lists at scale. Each also produces errors of its own, and the ones that matter here are errors of relevance: a protein can be on the list without the literature saying it does anything in the disease.

Biomedical natural language processing (NLP) now runs mostly on transformer language models pre-trained on biomedical text. PubMedBERT (Gu et al, 2022), BioBERT (Lee et al, 2020) and SciBERT (Beltagy et al, 2019) are the standard starting points for entity recognition, relation extraction and document classification. They differ mainly in pre-training corpus and vocabulary. PubMedBERT, trained from scratch on PubMed rather than continued from a general-domain model, scores highest on PubMed-derived tasks and beats general-domain Bidirectional Encoder Representations from Transformers (BERT) (Devlin et al, 2019).

CaseOLAP (Case-based Online Analytical Processing) (Liem et al, 2018, 2019; Fang et al, 2021, 2023; Santana et al, 2026) scores entities such as proteins, drugs and biomarkers against disease categories by how often they co-occur across large document collections. It has been used for extracellular matrix proteins in cardiovascular disease (Liem et al, 2018), organellar pathways through knowledge graphs (Fang et al, 2023), oxidative stress and calcium-regulating proteins (Bhatt et al, 2024), and maternal proteins in preeclampsia (Urdang et al, 2025). But co-occurrence cannot tell a finding from a mention. A sentence reporting that *“49% of HFpEF patients [were] on ACE inhibitors”* puts ACE next to HFpEF and says nothing about what ACE does in HFpEF. When we inspected CaseOLAP’s HFpEF output by hand, many high-scoring proteins were supported by sentences like this one.

We add a filter after CaseOLAP. For every PubMed sentence that names a protein and an HFpEF term, a classifier decides whether the sentence asserts a biological association, explicitly denies one, or mentions the protein incidentally. The classifier is a PubMedBERT model fine-tuned on 1,102 expert-labeled sentences with R-Drop and class-weighted cross-entropy, run as a six-model majority-vote ensemble over three seeds. Its labels then filter and re-weight CaseOLAP’s ranking, so that a protein rises because sentences say it matters, not because it is mentioned often. The paper contributes the classifier and ensemble; an audit method that uses ensemble disagreement to find mislabeled sentences, which surfaced 162 corrections; and an account of where that label work sat among the architecture, loss and threshold changes we tried.

Section 2 covers related work. Section 3 describes the data, annotation, model and audit. Section 4 reports the development chronology, seed stability and audit outcomes, and Section 5 the final results, errors and effect on CaseOLAP. Section 6 discusses what the label audit can and cannot show, and Section 7 concludes.

## 2 Related Work

### 2.1 Biomedical Relation Extraction

The three common biomedical encoders differ in what they were trained on. BioBERT (Lee et al, 2020) starts from general-domain BERT (Devlin et al, 2019) and continues pre-training on PubMed abstracts and PMC full text, so it keeps BERT’s general vocabulary. SciBERT (Beltagy et al, 2019) is trained from scratch on 1.14M Semantic Scholar papers, mostly computer science and biomedicine, with a vocabulary built from that corpus. PubMedBERT (Gu et al, 2022) is trained from scratch on PubMed alone, with a PubMed vocabulary, so rare biomedical terms are not broken into many sub-word pieces; it outperforms BioBERT and SciBERT on the BLURB benchmark. We use PubMedBERT for that reason.

Sentence-level biomedical relation extraction has dedicated corpora. ChemProt (Peng et al, 2018) labels chemical–protein interactions (inhibitor, agonist, substrate, antagonist) in PubMed abstracts. The DDIExtraction shared task (Segura-Bedmar et al, 2013) covers drug–drug interactions in DrugBank and MEDLINE text. BioRED (Luo et al, 2022) annotates 600 PubMed abstracts with relations among genes, diseases, chemicals and variants, including positive, negative and association-strength labels. EU-ADR (van Mulligen et al, 2012) annotates 300 abstracts for adverse drug reactions and drug–target relations. All of these type relations between entities. None separates a sentence that asserts a biological association from one that merely mentions both entities, and that separation is our task.

### 2.2 Label Noise in NLP

Expert-annotated datasets contain label errors, and those errors hurt classifiers (Northcutt et al, 2021; Song et al, 2022). One family of methods tolerates noise during training through robust losses, noise-aware weighting and sample re-weighting; Song et al (2022) survey it. The other finds the bad labels and fixes them. Confident Learning (Northcutt et al, 2021) estimates which labels are wrong from out-of-fold predicted probabilities and class-wise confidence thresholds, then prunes or relabels them; the Cleanlab library implements it for vision, text and tabular data.

Our setting is different in two ways. The dataset is small (1,402 sentences), so a domain expert can review every flagged sentence. And one annotator produced the labels, with no calibration against other raters. We therefore use a simpler signal than Confident Learning: train an ensemble across seeds and regularization settings, flag sentences where the majority vote disagrees with the label, rank them by disagreement rate, confidence in the wrong label and predictive entropy, and give the queue to an expert who sees the full abstract and the target protein. Section 3.3 gives the procedure.

### 2.3 CaseOLAP and Knowledge Extraction

CaseOLAP (Liem et al, 2018, 2019) scores entity–category associations by textual co-occurrence. CaseOLAP LIFT (Fang et al, 2021) adds knowledge-graph construction for protein–disease associations, and later work applied the platform to organellar pathways (Fang et al, 2023), oxidative stress (Bhatt et al, 2024) and preeclampsia (Urdang et al, 2025). Our classifier sits downstream of CaseOLAP and judges each co-occurrence at the sentence level (Figure 1).

**Fig. 1:**
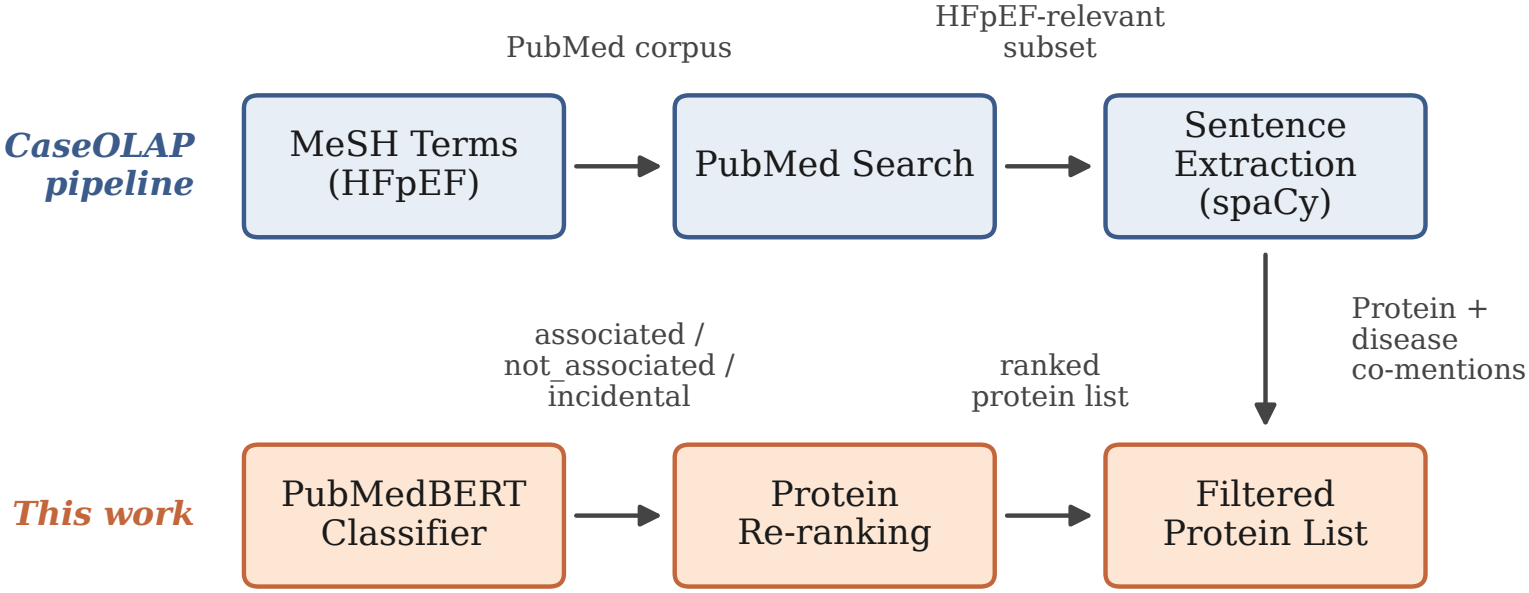
Pipeline. CaseOLAP retrieves abstracts and extracts protein mentions. The classifier (red) labels each mention associated, not_associated or incidental, and the re-ranking step orders proteins by association evidence instead of co-occurrence frequency.

## 3 Methods

### 3.1 Data Collection and Annotation

We retrieved PubMed abstracts through the CaseOLAP pipeline, which queries the National Library of Medicine’s E-utilities API with a Boolean expression of HFpEF-related Medical Subject Headings (MeSH) descriptors and free-text synonyms. The MeSH descriptors came from the C14 (cardiovascular diseases) branch, mainly C14.280.067, C14.280.238, C14.280.434, C14.280.484 and C14.280.647 (heart failure and its subtypes), together with the keywords “heart failure with preserved ejection fraction”, “HFpEF”, “diastolic heart failure” and “preserved ejection fraction”. Protein names and synonyms came from UniProt human entries. The full MeSH list and protein synonym table are in the supplementary material. Abstracts were split into sentences with spaCy, and every sentence containing at least one protein mention and one disease mention became a classification candidate.

A domain expert labeled each sentence with one of three classes:

- associated (32.1%): the sentence asserts a biological relationship between the protein and the disease.
- not_associated (23.8%): the sentence denies a relationship or reports a negative finding.
- incidental (44.1%): the sentence mentions the protein without asserting or denying a relationship.

After leak removal the training set has 1,102 sentences. The evaluation set has 300: 115 associated, 79 not_associated and 106 incidental. Table 1 gives examples, including a boundary case.

**Table 1.** Example sentences for each class. The boundary case shows the associated/incidental ambiguity that makes annotation hard.

| **Label** | **Sentence** |
|---|---|
| associated | “Higher levels of ADMA and SDMA and lower levels of homoarginine are associated with an adverse cardiovascular risk profile and diastolic dysfunction.” |
| associated | “Significant correlation between plasma Galectin-3 and E/Em in advanced HFPEF patients was noted.” |
| not_associated | “Lp(a) ≥50 mg/dL was not associated with HF risk overall or by EF subtype (HFpEF HR, 0.97).” |
| not_associated | “sST2 was not associated with left ventricular structure or left ventricular systolic or diastolic function.” |
| incidental | “In the HFpEF group, 76% were on beta-blockers, 49% on ACEi/ARB, 18% on ARNI, 49% on MRA and 40% on SGLT2i.” |
| incidental | “Our study aimed to differentiate patients with ATTRwt-CM from ATTRwt-negative HFpEF patients by identifying circulating protein biomarkers.” |
| *Boundary* | “HFpEF and HFrEF share elevated levels of leptin and adiponectin.” *(Labeled associated because it reports a finding. A reader who treats shared elevation as background, not a claim, would call it incidental.)* |

### 3.2 Model Architecture

We fine-tune PubMedBERT (Gu et al, 2022) (110M parameters) for three-way classification, with a linear head on the `[CLS]` representation.

#### Final training loss

The final v9 retraining stage uses class-weighted cross-entropy, with weights inversely proportional to class frequency (Eq. (1)):

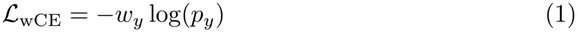

where *p*_*y*_ is the predicted probability of the gold label *y* and *w*_*y*_ is its class weight.

#### Earlier focal-loss baseline

Earlier ablations and the v7 base checkpoint used focal loss (Lin et al, 2017) with *γ* = 2.0 (Eq. (2)):

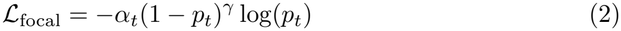

where *p*_*t*_ is the predicted probability of the true class and *α*_*t*_ is the class weight.

#### R-Drop regularization

Fine-tuning 110M parameters on about 1,100 examples gives results that swing with the random seed. R-Drop (Wu et al, 2021) damps this by passing each example through the model twice with different dropout masks and penalizing disagreement between the two output distributions (Eq. (3)):

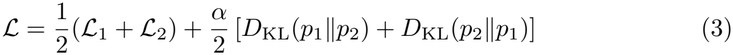

with *α* = 0.5.

#### Threshold tuning, reported as sensitivity analysis only

We also tried temperature scaling (Guo et al, 2017) and per-class threshold search with a minimum recall constraint on associated. Earlier drafts tuned those thresholds on the evaluation set itself, which flatters the result, so we report those numbers only as sensitivity analysis. Headline results use the uncalibrated majority vote and touch the evaluation set once; the uncalibrated cross-seed mean (84.4 *±*0.7% accuracy) is the threshold-free reference.

#### Ensemble

We train six models, three with R-Drop (*α* = 0.5) and three with standard dropout, over seeds 42, 2026 and 314. Headline predictions are their majority vote.

### 3.3 Label Audit

The audit treats ensemble disagreement as a pointer to possible annotation errors:

1. Train *k* = 6 models with different seeds and regularization settings.
2. Collect all *k* predictions for each evaluation example.
3. Flag examples whose majority vote differs from the current label.
4. Rank the flagged examples by a suspicion score combining disagreement rate, confidence in the wrong label and predictive entropy.
5. Show each one to a domain expert with the original abstract and the target protein.
6. Apply the corrections and retrain.

We ran two rounds. In the first, the majority vote disagreed with the label on 88 of 300 evaluation examples (29.3%). Expert review corrected 53 of them and exposed a flaw in the annotation sheet: when a sentence named several proteins, the sheet did not say which protein the label was for. We added a *target protein* column for later rounds. The second round ran the same procedure on 164 training examples and corrected 109.

### 3.4 Evaluation Protocol

We report accuracy and macro-averaged F1 on the 300-sentence evaluation set. Macro F1 weights the three classes equally, so a model cannot score well by ignoring the smaller not_associated class. Cross-seed statistics (mean *±* standard deviation) are over all six models. As an exploratory transfer check, we also evaluate on EU-ADR (van Mulligen et al, 2012) and BioRED (Luo et al, 2022) after mapping their labels to our task.

## 4 Experiments

### 4.1 Experimental Setup

All models start from the PubMedBERT checkpoint `microsoft/BiomedNLP-PubMedBERT-base-uncased-abstract-fulltext` (110M parameters). Hyperparameters: learning rate 2 *×* 10^*−*5^, batch size 16, at most 6 epochs with early stopping (patience 2), weight decay 0.01. Models are warm-started from a base checkpoint trained on the relabel3 version of the same 1,102-example HFpEF split (v7). Training ran on an AWS EC2 r8g.2xlarge instance (8-core ARM Graviton, 64 GB RAM, CPU only), about 50 minutes per model.

#### Reproducibility details

Each run used a 0.15 validation split, maximum sequence length 256, and best-checkpoint selection by validation weighted F1. The final v9 runs were warm-started from `models/hfpef_v7/pubmedbert_focal_ large_base_20260224/final`; that checkpoint was trained with focal loss, while the v9 stage itself used weighted cross-entropy. Temperature scaling searched *T ∈ {*0.7, 0.8, 0.9, 1.0, 1.1, 1.25, 1.5, 2.0*}*. Per-class thresholds were searched over associated *{*0.30, 0.35, 0.40, 0.45*}*, not_associated *{*0.45, 0.50, 0.55, 0.60*}* and incidental *{*0.25, 0.30, 0.35, 0.40*}*, with a minimum associated recall of 0.45.

### 4.2 Development Chronology

Table 2 lists the model and data changes in the order we made them.

**Table 2.** Development chronology. The first row is the co-occurrence baseline, which labels every evaluation sentence as associated. The other rows are model milestones on three evaluation sets: the original 176 sentences, the 300-sentence relabel3 set and the final 300-sentence relabel4 set. Both the model and the ground truth change between groups, so read the rows as history, not as a controlled same-set ablation.

| **Configuration** | **Eval set** | **Acc. (%)** | **Macro F1 (%)** |
|---|---|---|---|
| CaseOLAP co-occurrence (all associated) | 300 / relabel4 | 38.3 | 18.5 |
| Sentence-only PubMedBERT (focal-loss baseline) | 176 / orig. | 69.3 | 66.7 |
| + Threshold-searched fusion system | 176 / orig. | 72.2 | 69.7 |
| + Calibrated v7 base model | 300 / relabel3 | 77.7 | 76.9 |
| + Relabel4 audit + weighted CE retrain | 300 / relabel4 | 85.3 | 85.9 |
| + Eval-tuned calibrated single | 300 / relabel4 | 86.3 | 87.1 |
| + 6-model vote ensemble | 300 / relabel4 | **85.3** | **86.1** |

*Sensitivity analysis.* With thresholds tuned on the evaluation set, the same six-model ensemble reaches 87.3% accuracy and 88.0% macro F1.

The post-audit row in Table 2 is the best single relabel4 model. Figure 2 plots the same history with the uncalibrated cross-seed mean at that stage (84.4%) instead, so the gain from ensembling stays visible. The rise from 76.9% macro F1 (relabel3, calibrated v7) to 86.1% (relabel4 ensemble) is not a same-set gain: the earlier number is scored against the relabel3 labels and the later one against the relabel4 set, in which 53 labels were corrected. It mixes label correction, the relabeling pass and later modeling changes.

**Fig. 2:**
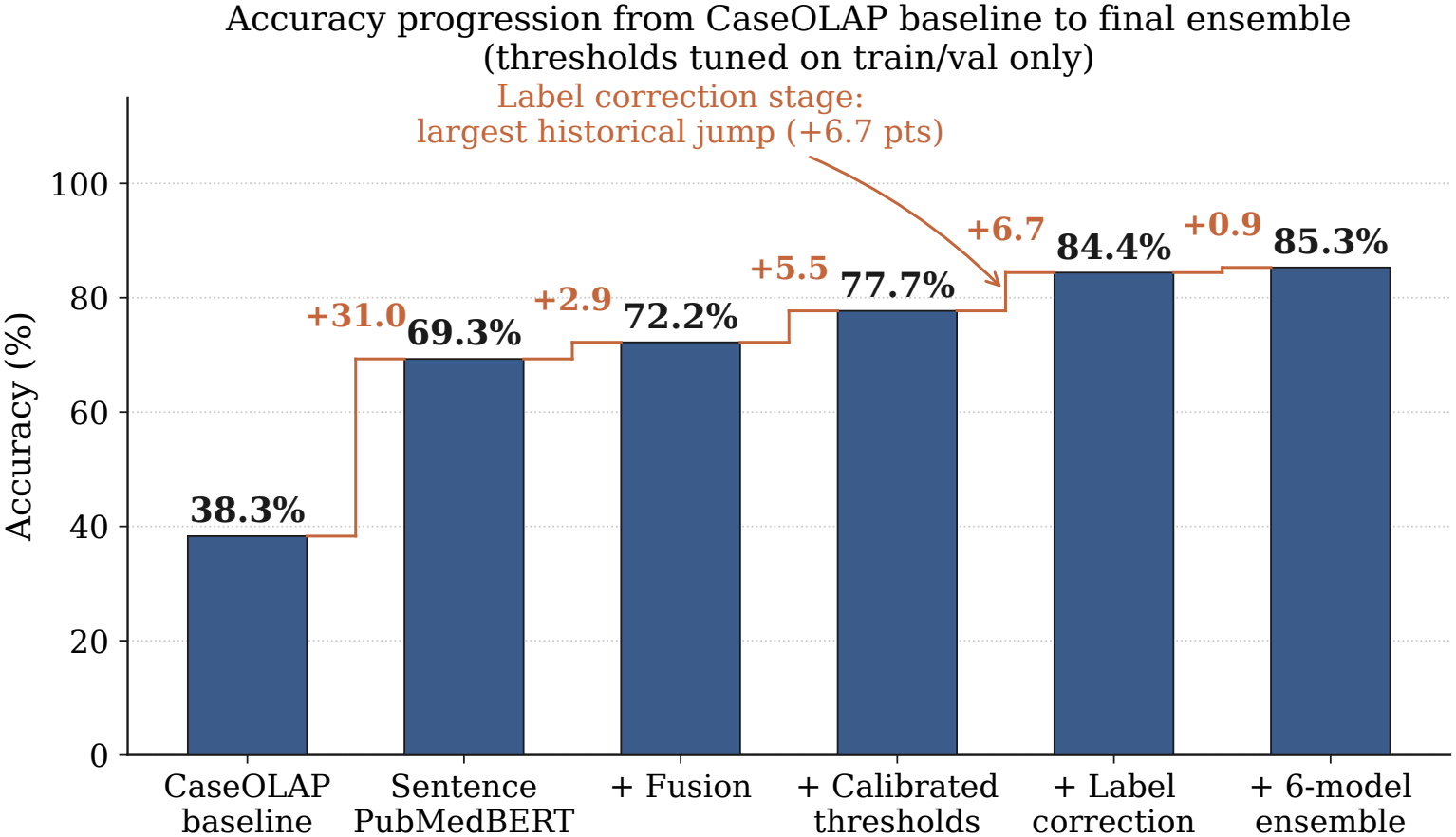
Accuracy across development milestones. The relabel4 retrain is plotted as the uncalibrated cross-seed mean of single models (84.4%); the last point is the six-model ensemble (85.3%).

### 4.3 Seed Stability

Across the six single models (three R-Drop, three standard dropout), uncalibrated accuracy was 84.4 *±*0.7% and macro F1 85.2 *±*0.6%. With per-run calibrated thresholds, accuracy was 83.9*±*0.7% and macro F1 84.8*±*0.7% (`logs/v9_train_calibrated_eval.json`). A standard deviation under one percentage point means run-to-run variance is modest.

### 4.4 Label Audit Results

Table 3 breaks down the training corrections by direction.

**Table 3.** Training-label corrections by direction in the second audit round. The first round, on the evaluation set, corrected 53 more. The most common change, incidental *→*associated, came from multi-protein sentences.

| **Correction direction** | **Count** |
|---|---|
| incidental → associated | 57 |
| associated → incidental | 27 |
| associated → not_associated | 10 |
| incidental → not_associated | 9 |
| not_associated → incidental | 4 |
| not_associated → associated | 2 |
| Total training corrections | 109 |
| Total evaluation corrections (round 1) | 53 |

More than half of the training corrections (57 of 109, 52.3%) moved a sentence from incidental to associated. These sentences named several proteins. The annotator had labeled a different protein in the sentence, one that really was incidental, while the target protein was associated with the disease.

## 5 Results

### 5.1 Main Results

Table 4 gives per-class results for the best uncalibrated single model and the six-model majority vote, with the cross-seed mean of the single models. not_associated is the easiest class (F1 93.8%): negative findings usually come with explicit phrases such as “did not differ” and “did not correlate.” The hard line is between associated and incidental, where the majority vote scores F1 82.5 and 81.9.

**Table 4.** Main results on the 300-sentence evaluation set (115 associated, 79 not_associated, 106 incidental), with no thresholds tuned on it.

| System | Class / setting | Prec. | Rec. | F1 | Acc. |
|---|---|---|---|---|---|
| Best single (uncalibrated) | associated | 88.8 | 75.7 | 81.7 | – |
|  | not_associated | 89.4 | 96.2 | 92.7 | – |
|  | incidental | 79.5 | 87.7 | 83.4 | – |
|  | *Macro avg* | 85.9 | 86.5 | 85.9 | 85.3 |
| 6-model majority vote | associated | 85.2 | 80.0 | 82.5 | – |
|  | not_associated | 91.6 | 96.2 | 93.8 | – |
|  | incidental | 80.7 | 83.0 | 81.9 | – |
|  | *Macro avg* | 85.8 | 86.4 | 86.1 | 85.3 |
| Cross-seed mean | Uncalibrated single models | – | – | 85.2 | 84.4 |

*Sensitivity analysis.* With thresholds tuned on the evaluation set, the best single model reaches 86.3% accuracy and 87.1% macro F1, and the six-model ensemble 87.3% and 88.0%.

Figure 3 shows the confusion matrices before and after label correction. The change is concentrated in the associated/incidental cells: correction reduces the earlier underprediction of associated, and the remaining errors still sit on that boundary, not in not_associated.

**Fig. 3:**
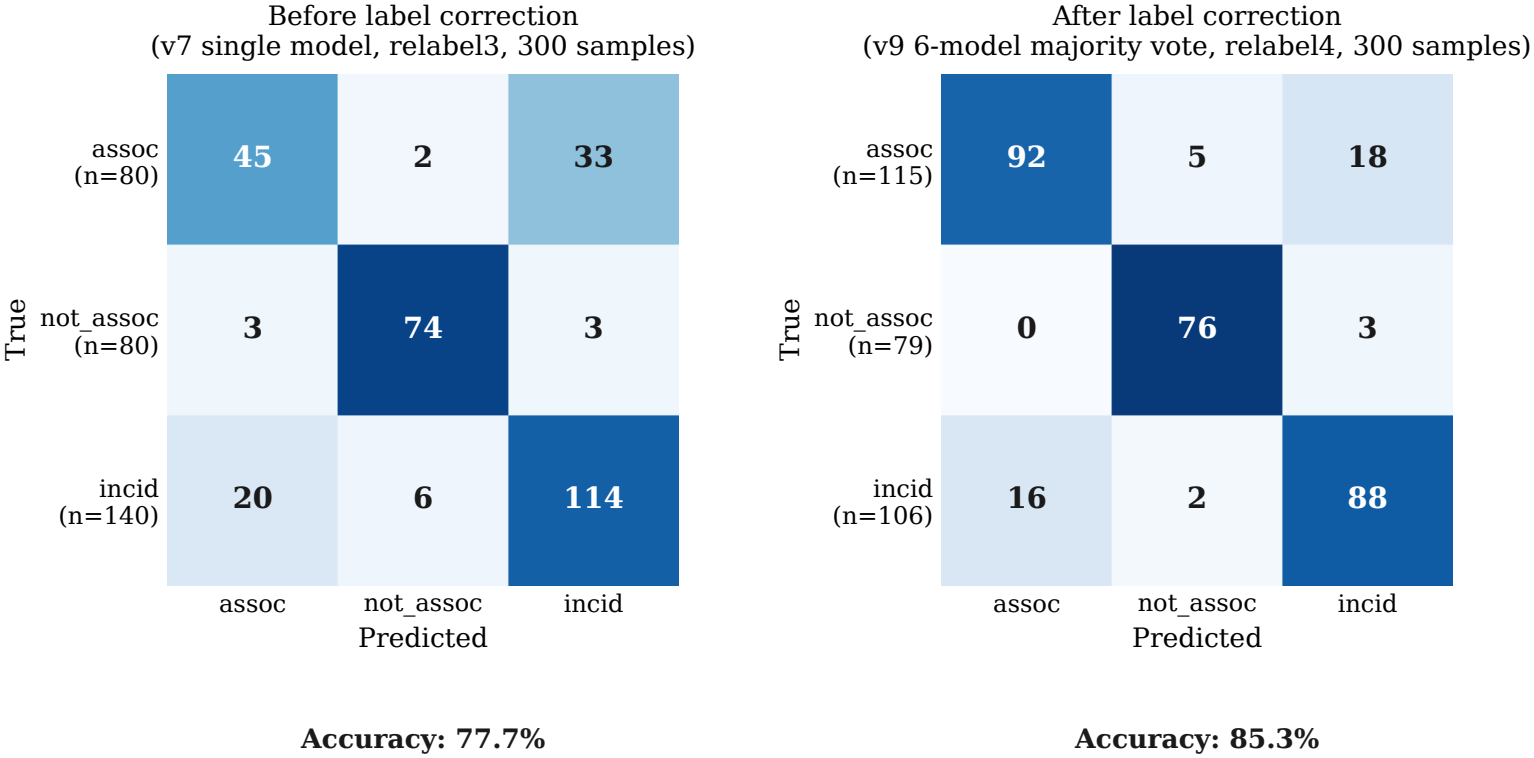
Confusion matrices before and after label correction. Left: v7 single model with the best constrained thresholds on the relabel3 300-sentence evaluation set (`hfpef_v7_large_base_calibration_20260224.json`). Right: v9 six-model majority vote on the relabel4 300-sentence evaluation set (`v9_publication_eval.json`). Rows are normalized. The associated/incidental cells shift most.

### 5.2 Error Analysis

The majority vote makes 44 errors on the 300 sentences: 18 associated *→* incidental (40.9%), 16 incidental *→* associated (36.4%), 5 associated *→* not_associated (11.4%), 3 not_associated *→* incidental (6.8%) and 2 incidental *→* not_associated (4.5%). Thirty-four of the 44 (77.3%) are on the associated/incidental boundary.

We see three recurring patterns. When a finding is compressed into terse clinical prose, the model backs off to incidental: “The severity of cardiac fibrosis evaluated by CMRI also correlated with CTGF.” When a sentence pairs a protein with a positive relation word but never ties the protein to the disease, the model over-calls associated: “Notably, poor nutritional status was significantly associated with GDF15.” And sentences that mix a null comparison with a positive one remain hard: “In men, adjusted similarly as leptin, there was no difference between HFpEF and HFrEF, p=0.310 but, compared to controls, higher levels in HFpEF (p=0.044) and HFrEF (p=0.001)” is labeled associated and predicted not_associated.

### 5.3 Effect on CaseOLAP Protein Rankings

Run after CaseOLAP, the classifier changes the protein ranking, because proteins supported mostly by incidental or negative sentences lose evidence. The ranking moves away from how often a protein is mentioned and toward how often a sentence says it is involved. This paper benchmarks that filtering step at the sentence level; the exact protein-level ordering depends on the particular CaseOLAP export and identifier normalization and is left to downstream analysis.

### 5.4 Exploratory Transfer Evaluation

We also ran the classifier on two external datasets after mapping their labels to ours (Table 5). Neither task matches ours, so these are transfer checks, not benchmarks.

**Table 5.** Exploratory transfer evaluation after task-specific label mapping.

| **Dataset** | **Acc. (%)** | **Caveat** |
|---|---|---|
| EU-ADR | 82.9 | 2,891 relations; labels mapped to a binary associated/incidental scheme by merging not_associated into associated. |
| BioRED | 94.0 | Random 200-relation gene–disease subsample, run through the end-to-end extraction pipeline rather than the standalone classifier; only 11 gold not_associated cases. |

EU-ADR reached 82.9% accuracy, but the binary remapping makes it an easier task than our three-way one. BioRED reached 94.0%, but 189 of the 200 relations were positive after remapping, and the pipeline got none of the 11 negatives right.

## 6 Discussion

### 6.1 What the Label Audit Shows

The biggest jump in the project came between the relabel3 calibrated v7 model (76.9% macro F1) and the relabel4 retrain (85.9% macro F1, single model). Three things changed at once: labels flagged by ensemble disagreement were corrected, a relabeling pass added the target-protein column (Section 3.3), and the retrain replaced focal loss with weighted cross-entropy. The evaluation set had already grown from 176 to 300 sentences before the relabel3 milestone (Table 2). We cannot separate the three changes with the runs we have. Claiming that label correction caused the gain would take a same-set test that fixes the gold standard before and after correction.

Within the final relabel4 setting, the later changes add little. The audited single model reaches 85.9% macro F1 and the six-model vote 86.1%, a gain of 0.2 points. We do not rank these steps against the audit: they happened at different stages, on different evaluation and label sets.

Label noise is known to hurt classifiers (Northcutt et al, 2021; Song et al, 2022). Here, the largest single source of corrected labels was one flaw in the annotation interface. Whether that flaw is the main source of label noise in this corpus, or one of several, a single audit cannot tell.

### 6.2 Multi-Protein Sentences

Biomedical sentences often name several proteins. An annotator asked to “classify the protein–disease relationship in this sentence” has to know which protein the question means, and our first annotation sheet did not say. Of the 109 training corrections, 57 were sentences of this kind. The target-protein column reduced the ambiguity in later rounds, though we did not run a comparison that varies only the sheet design.

Naming the target protein does not settle every case. “Galectin-3 was measured at baseline” states no relationship, yet a reader who sees galectin-3 measured in study after study may take that as weak evidence of association. Our labels encode one expert’s standard. Measuring how often reasonable readers disagree on such cases would take several independent annotators.

### 6.3 Ensemble Disagreement as an Audit Signal

The six-model vote disagreed with the label on 88 of 300 evaluation examples (29.3%), and expert review corrected 53 of them (60.2%). The method needs no probability calibration and works with any set of models. It trades recall for precision: it sends the reviewer only the sentences where the ensemble leans away from the label, so mislabeled sentences with low-margin predictions go unreviewed. Confident Learning (Northcutt et al, 2021) catches more, at the cost of requiring calibrated out-of-fold probabilities.

### 6.4 Limitations

1. Thresholds tuned on the evaluation set appear only as sensitivity analysis and are optimistic; headline results use the uncalibrated majority vote on an untouched evaluation set.
2. Label correction changed the evaluation set between the before and after comparisons, so the chronology table compares models against different ground truth.
3. One domain expert reviewed all labels, and we did not measure inter-annotator agreement (Cohen’s *κ*), so annotation subjectivity is unquantified. The open question is the associated/incidental boundary, as in the galectin-3 example above. Independent annotators on a stratified subset would show how often that boundary changes the corrected labels and the metrics.
4. A training set of 1,102 sentences limits generalization. In the BioRED transfer check, not_associated recall was 0% on the 11 negative examples.
5. With 300 evaluation sentences the bootstrap intervals are wide: 81.0–89.3% for accuracy and 82.1–89.7% for macro F1 for the majority vote.
6. The only baseline on the relabel4 evaluation set is the all-associated co-occurrence baseline (38.3% accuracy). We did not run a TF-IDF logistic regression, a lexical-rule classifier or a calibrated zero-shot LLM on the same split.

## 7 Conclusion

Most sentences that put a protein next to HFpEF do not say the protein is involved in it. A six-model PubMedBERT ensemble separates the ones that do, with 85.3% accuracy and 86.1% macro F1 on 300 held-out sentences, against 38.3% for co-occurrence. The largest gain in the project came at the stage where we audited labels and named the target protein in multi-protein sentences; because that stage changed other things too, we report it as history, not proof. Our experience suggests that, for small expert-labeled biomedical datasets, the annotation sheet and the labels deserve as much engineering attention as the model.

## Data Availability

The source sentences come from publicly available PubMed abstracts. The corrected training and evaluation datasets, trained model checkpoints and evaluation scripts produced in this study are available from the corresponding author on reasonable request, pending institutional review.

## Acknowledgments

This work was conducted in the Division of Cardiovascular Medicine, Department of Internal Medicine, University of California, Davis. Computational resources were provided by Amazon Web Services.

## Data Availability

The corrected training and evaluation datasets, trained model check-points and evaluation scripts are available on request, pending institutional review. In the project repository, the relabel4 training set is `data/hfpef_v9_train_autocorrect109.json`, the 300-sentence evaluation set is `data/splits/hfpef_v8_eval_relabel4_noleak_large.json`, the publication results are `logs/v9_publication_eval.json`, and the calibrated cross-seed results are `logs/v9_train_calibrated_eval.json`. Protein synonyms for the CaseOLAP retrieval query came from UniProt human entries through the CaseOLAP pipeline; the exact synonym table is in the supplementary material.

## Declarations

### Ethics

This study analyzed sentences from published PubMed abstracts only. It involved no human participants, patient records or animals, so institutional review board approval was not required.

### Author contributions

All authors contributed to data analysis, interpretation of the findings, manuscript preparation, and critical revision of the intellectual content. All authors reviewed and approved the final manuscript and agreed to be accountable for the integrity of the work.

